# Klebsiella pneumoniae infection increases risk of Alzheimer’s Disease in the UK Biobank cohort

**DOI:** 10.1101/2024.11.21.24317739

**Authors:** Steven Lehrer, Peter H. Rheinstein

## Abstract

**Background:** Infections, including bacterial pathogens, have been implicated in Alzheimer’s disease (AD) risk. Klebsiella pneumoniae (K. pneumoniae) is a common hospital-acquired pathogen associated with significant inflammation, which may contribute to neurodegeneration. This study investigates the relationship between K. pneumoniae infections and AD in the UK Biobank cohort.

**Methods:** Using UK Biobank data, we assessed AD diagnoses based on linked healthcare records and identified K. pneumoniae infections using ICD-10 codes B96.1 and J15.0. A cohort of 502,494 participants was analyzed for AD incidence in relation to demographic factors, educational years, APOE isoforms, and history of K. pneumoniae infection. Logistic regression was used to assess the association between K. pneumoniae infection and AD risk.

**Results:** AD incidence was significantly higher among participants with a history of K. pneumoniae infection (1.0%) compared to those without (0.2%; p < 0.001, Fisher’s exact test two tailed). Logistic regression analysis revealed that K. pneumoniae infection was associated with an increased risk of AD (OR = 3.32, p < 0.001), independent of age, sex, education, and APOE isoform. Additionally, AD risk was higher among ε4ε4 carriers and increased with age but decreased with additional years of education.

**Conclusion:** Our findings suggest that K. pneumoniae infection may be an independent risk factor for AD. This association underscores the need for further research into infection control and its role in mitigating neurodegenerative disease risk, particularly in populations susceptible to healthcare-associated infections.

**Graphical abstract:** Klebsiella pneumoniae infection increases risk of Alzheimer’s Disease. The gut-brain axis may be involved.

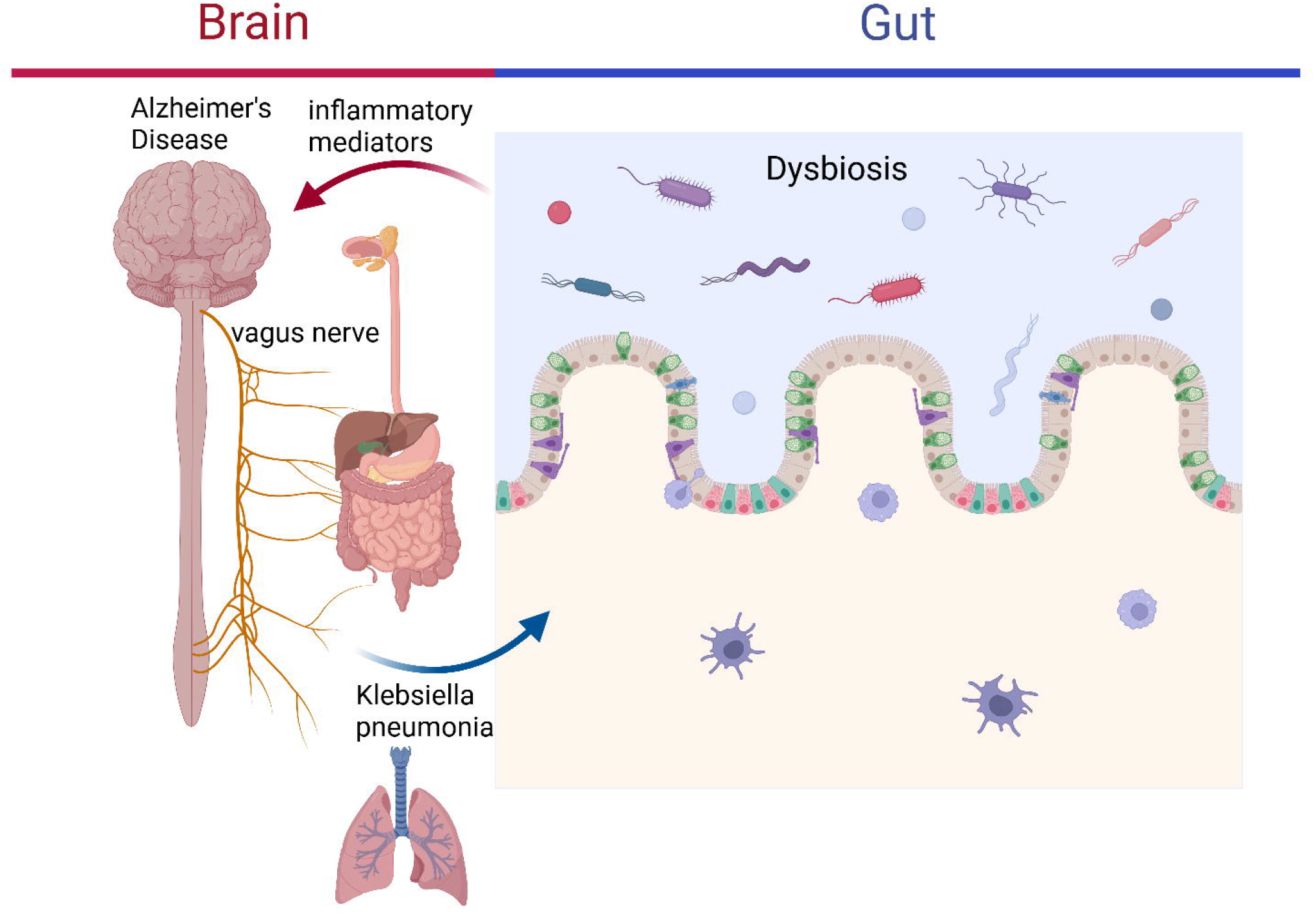

---

Park et al investigated the effects of Klebsiella pneumoniae (K. pneumoniae) infection on Alzheimer’s Disease (AD) pathology, using a 3xTg-AD mouse model. They found that K. pneumoniae, especially under antibiotic-induced dysbiosis, can breach the gut barrier, enter the bloodstream, and infiltrate the brain, causing neuroinflammation and impairing neurobehavioral functions. Dysbiosis is an imbalance between the types of organism present in natural human microflora, especially in the gut, that may contribute to many illnesses. Mice infected with K. pneumoniae showed increased levels of pro-inflammatory cytokines (e.g., IL-1β, IL-6, IL-8, TNF-α) in brain regions like the hippocampus and frontal cortex, which correlated with neurobehavioral impairments in tasks assessing memory and motor function. Antibiotic treatment exacerbated K. pneumoniae colonization and spread, highlighting the role of dysbiosis in enabling infection and neuroinflammatory response. The study by Park et al supports a possible link between infections, gut dysbiosis, and AD pathology, suggesting that health care-associated infections and antibiotic use in vulnerable populations may accelerate neurodegeneration through the gut-brain axis [1].

Infections have been linked to an increased risk of Alzheimer’s disease [2-5]. Viruses like herpes simplex virus (HSV-1) and varicella-zoster virus (VZV) have been associated with Alzheimer’s disease [6]. These viruses can cause chronic inflammation in the brain, which may contribute to the development of Alzheimer’s. Infections can lead to long-term activation of the immune system, which may increase the risk of neurodegenerative diseases. Persistent infections can cause chronic inflammation, which is a known risk factor for Alzheimer’s disease. People with Alzheimer’s disease often have a weakened immune system, making them more susceptible to infections. While the exact mechanisms are still being studied, it’s clear that infections play a role in the complex process of Alzheimer’s disease development [7].

In the current study we used UK Biobank data to assess the relationship of K. pneumoniae to AD.

## Methods

The UK Biobank is a large-scale biomedical database and research resource that collects in-depth genetic, lifestyle, and health information from about 500,000 volunteer participants across the United Kingdom. Launched in 2006, it aims to support research into the causes, prevention, and treatment of a wide range of diseases, including cancer, cardiovascular disease, diabetes, Alzheimer’s, and mental health disorders.

UK Biobank identifies Alzheimer’s disease in participants primarily through linked healthcare records, including hospital inpatient and outpatient records, death registries, and primary care data. Diagnoses are coded based on ICD (International Classification of Diseases) codes, allowing researchers to identify Alzheimer’s disease cases using both ICD-9 and ICD-10 codes, as well as dementia-related codes that may capture Alzheimer’s-related diagnoses. The most used codes include:

- **ICD-10 code** G30 (for Alzheimer’s disease), with subcategories G30.0 (early onset), G30.1 (late onset), and G30.9 (unspecified).
- **ICD-9 code** 331.0, which corresponds to Alzheimer’s disease.

Additionally, the UK Biobank captures dementia-related information through cognitive tests conducted during assessment visits. These include tests of reaction time, memory, and fluid intelligence, among others, which may reflect cognitive impairment trends associated with dementia, including Alzheimer’s disease. Genetic information is available for many participants, allowing for analyses related to Alzheimer’s disease risk, including the APOE ε4 gene isoform known to influence Alzheimer’s risk.

UK Biobank identifies APOE isoforms (specifically APOE ε2, ε3, and ε4) through genotyping and imputation. They assess specific single nucleotide polymorphisms (SNPs) associated with the APOE gene that determine an individual’s isoform. The SNPs used to identify APOE alleles are rs429358 and rs7412. These SNPs define the APOE genotype as follows:

- ε2 (Cys130Cys158): Characterized by the combination of rs429358-T and rs7412-T.
- ε3 (Cys130Arg158): Characterized by rs429358-T and rs7412-C.
- ε4 (Arg130Arg158): Characterized by rs429358-C and rs7412-C.

Based on the combinations of these alleles, individuals are classified into one of the APOE genotypes, such as ε2/ε2, ε2/ε3, ε3/ε3, ε3/ε4, and ε4/ε4.

UK Biobank does not routinely test participants directly for Klebsiella pneumoniae infections. However, it identifies participants who may have had Klebsiella pneumoniae infections by leveraging linked healthcare records. Infections can be identified through ICD (International Classification of Diseases) codes in hospital and primary care records, which indicate diagnosis and treatment of bacterial infections, including those caused by Klebsiella species. Relevant ICD-10 codes include:

ICD-10 code B96.1: *Klebsiella pneumoniae as the cause of diseases classified elsewhere*, which identifies cases where Klebsiella pneumoniae has been confirmed as the causative agent of infection.

ICD-10 code J15.0 pneumonia due to Klebsiella pneumoniae.

UK Biobank links participants’ records to microbiological laboratory data where available.

In the UK, hospital laboratories routinely perform cultures and sensitivity testing on respiratory or blood samples to confirm the presence of bacterial pathogens like Klebsiella pneumoniae. When these records are linked, they can provide more detailed insights into specific infections and antimicrobial resistance.

Statistics were computed with SPSS v 26 (IBM, New York).

## Results

Table 1 shows demographics of sample.

**Table 1.** Demographics of subjects in this study.

|  |  |  |  |
| --- | --- | --- | --- |
|  | Alzheimer's Disease |  |  |
| 460 females | 48.30% | 493 males | 51.70% |
| 96% white British |  |  |  |
| age 56 ± 8 (mean ± |  |  |  |
| SD) |  |  |  |
| years education 15 ± 5 |  |  |  |
|  | Klebsiella pneumonia |  |  |
| 610 females | 42.50% | 826 males | 57.50% |
| 92% white British |  |  |  |
| age 56 ± 8 (mean ± SD) |  |  |  |
| years education 15 ± 5 |  |  |  |

Table 2 shows Alzheimer’s disease (AD) versus K. pneumoniae infection in 502,494 subjects. In subjects with AD 0.2% had not had K. pneumoniae infection. In subjects with AD 1.0% had K. pneumoniae infection. The effect of K. pneumoniae on AD incidence was significant (p < 0.001, two tailed Fisher exact test.)

**Table 2.** Alzheimer’s disease (AD) versus K. pneumoniae infection in 502,494 subjects. In subjects without AD 0.2% had not had K. pneumoniae infection. In subjects with AD 1.00% had K. pneumoniae infection. The effect of K. pneumoniae on AD was significant (p < 0.001, two tailed Fisher exact test.)

|  |  | K. pneumoniae | K. pneumoniae |  |
| --- | --- | --- | --- | --- |
| Alzheimer's |  | no | yes | total |
| no | Count | 500119 | 1422 | 501541 |
|  | % within K. pneumoniae | 99.80% | 99.00% | 99.80% |
| yes | Count | 939 | 14 | 953 |
|  | % within K. pneumoniae | 0.20% | 1.00% | 0.20% |
| total | Count | 501058 | 1436 | 502494 |
|  | % within K. pneumoniae | 100.00% | 100.00% | 100.00% |

Table 3 shows logistic regression with 95% confidence intervals, AD dependent variable; sex, age, years of education, ApoE isoform (ε3ε3 versus ε4ε4, ε3ε4 versus ε4ε4) K. pneumoniae infection (yes or no) dependent variables. AD was more common in men (Odds Ratio 1.151, p = 0.063). AD increased with each year of age (O.R. 1.213, p < 0.001). AD decreased with each additional year of education (O.R. 0.964, p < 0.001). AD was increased ε4ε4 versus ε3ε3 (O.R. 11.977, p < 0.001). AD was increased ε3ε4 versus ε3ε3 (O.R. 3.53, p < 0.001). K. pneumoniae infection increased risk of AD (O.R. 3.32, p < 0.001).

**Table 3.**
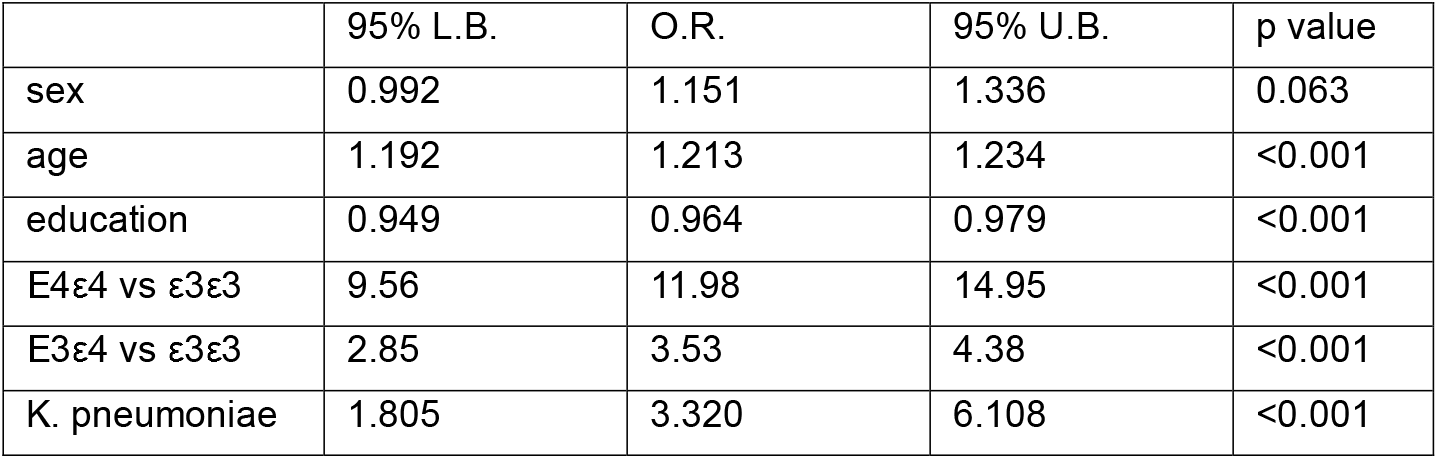
Logistic regression with 95% confidence intervals, AD dependent variable; sex, age, years of education, ApoE isoform (ε3ε3 versus ε4ε4, ε3ε4 versus ε4ε4) K. pneumoniae infection (yes or no) dependent variables. AD was more common in men (O.R. 1.151, p = 0.063). AD increased with each year of age (O.R. 1.213, p < 0.001). AD decreased with each additional year of education (O.R. 0.964, p < 0.001). AD was increased ε4ε4 versus ε3ε3 (O.R. 11.977, p < 0.001). AD was increased ε4ε4 versus ε3ε4 (O.R. 3.53, p < 0.001). K. pneumoniae infection increased risk of AD (O.R. 3.32, p < 0.001). L.B. lower bound, U.B. upper bound, O.R. odds ratio.

|  | 95% L.B. | O.R. | 95% U.B. | p value |
| --- | --- | --- | --- | --- |
| sex | 0.992 | 1.151 | 1.336 | 0.063 |
| age | 1.192 | 1.213 | 1.234 | <0.001 |
| education | 0.949 | 0.964 | 0.979 | <0.001 |
| E4ε4 vs ε3ε3 | 9.56 | 11.98 | 14.95 | <0.001 |
| E3ε4 vs ε3ε3 | 2.85 | 3.53 | 4.38 | <0.001 |
| K. pneumoniae | 1.805 | 3.320 | 6.108 | <0.001 |

In the UK Biobank Alzheimer’s disease is more common in men than in women [8, 9]. Several factors contribute to this difference:

Health-Related Conditions: Men in the study had higher rates of conditions like hypertension, diabetes, and cardiovascular diseases, which are known risk factors for dementia.

Biomarkers: Certain blood and urinary biomarkers, such as apolipoprotein A and creatinine levels, were more prevalent in men and contributed to the higher risk.

Lifestyle Factors: Men were more likely to engage in behaviors like smoking and had higher rates of unhealthy alcohol intake, both of which are associated with an increased risk of dementia.

Multimorbidity: Men had a higher multimorbidity risk score, which means they had more coexisting chronic conditions that could contribute to dementia.

## Discussion

*Klebsiella pneumoniae* is a Gram-negative, rod-shaped bacterium that is part of the *Enterobacteriaceae* family. While it is normally found in the human intestines and does not typically cause harm in healthy individuals, it can become pathogenic under certain conditions. This bacterium is a common cause of healthcare-associated infections, particularly in hospitals, where it can lead to serious infections, including pneumonia, urinary tract infections (UTIs), bloodstream infections, and wound or surgical site infections [10].

K. pneumoniae is rod-shaped and encapsulated, with a characteristic mucoid (slimy) appearance due to a thick polysaccharide capsule that protects it from immune defenses. It is facultatively anaerobic, meaning it can survive in both oxygen-rich and oxygen-poor environments. K. pneumoniae is known for its ability to acquire antibiotic resistance. Certain strains, particularly carbapenem-resistant *Klebsiella pneumoniae* (CRKP), are highly resistant to multiple antibiotics and pose a significant public health risk. *Klebsiella pneumoniae* infections are particularly dangerous in immunocompromised individuals or those with chronic illnesses, such as diabetes, liver disease, or lung disease [11].

K. pneumoniae capsule and other virulence factors, including lipopolysaccharide (LPS) and siderophores (molecules that scavenge iron), contribute to its ability to evade immune responses and establish infections. In cases of pneumonia, K. pneumoniae often leads to symptoms such as high fever, chills, productive cough with thick, blood-tinged sputum (“currant jelly” sputum), and chest pain. Its ability to cause severe, necrotizing lung infections means it can lead to complications like lung abscesses, cavitation, and empyema [12]. In the gut, K. pneumoniae, especially with antibiotic treatment, can be associated with dysbiosis.

Gut dysbiosis can involve the vagus nerve in Alzheimer’s disease. The vagus nerve is a key component of the gut-brain axis, a bidirectional communication system between the gut and the brain. Gut dysbiosis can lead to the production of inflammatory mediators and metabolites that can travel through the vagus nerve to the brain, contributing to neuroinflammation and the progression of Alzheimer’s disease [13].

Klebsiella pneumoniae infection could theoretically contribute to increased risk of Alzheimer’s disease through mechanisms involving chronic inflammation, immune system activation, and potential neuroinvasion. Here are several key pathways that might link K. pneumoniae to Alzheimer’s disease risk:

1. Chronic Systemic Inflammation: K. pneumoniae infections can lead to persistent or recurrent inflammation in the body, especially in immuno-compromised or elderly individuals. Chronic systemic inflammation is a known risk factor for Alzheimer’s disease, as inflammatory molecules (like cytokines) can cross the blood-brain barrier, potentially leading to neuroinflammation. This neuroinflammation is implicated in the progression of Alzheimer’s pathology, including amyloid-beta (Aβ) plaque deposition and tau protein hyperphosphorylation [1].
2. Blood-Brain Barrier (BBB) Disruption: Some studies suggest that infections can compromise the integrity of the BBB, allowing pathogens or inflammatory mediators to reach brain tissue. K. pneumoniae, particularly in cases of severe pneumonia or bloodstream infection (sepsis), could contribute to BBB disruption. This weakened barrier can expose the brain to systemic pathogens and inflammatory molecules that may promote Alzheimer’s pathology [1].
3. Direct Neuroinvasion: While rare, certain strains of K. pneumoniae, especially hypervirulent strains, have been implicated in meningitis and brain abscesses, indicating their potential to invade the central nervous system (CNS) [14]. If K. pneumoniae were able to reach brain tissue, it could trigger localized inflammation and oxidative stress, further contributing to the neurodegenerative processes involved in Alzheimer’s.
4. Molecular Mimicry and Immune System Activation: K. pneumoniae, like many pathogens, can produce antigens that resemble human proteins, leading to an autoimmune response [15]. This molecular mimicry could result in the immune system mistakenly attacking brain tissue or proteins like amyloid precursor protein (APP) or tau, accelerating Alzheimer’s-related damage.
5. Microbiome Interactions: K. pneumoniae is part of the human gut microbiome, and recent studies suggest that gut bacteria may influence brain health through the gut-brain axis [1]. Disruptions in gut microbiota composition such as an overgrowth of K. pneumoniae could contribute to systemic inflammation and alter immune responses in ways that affect brain function and increase Alzheimer’s risk. Elevated levels of certain gut bacteria have been associated with increased Aβ accumulation and cognitive decline, suggesting that K. pneumoniae might contribute in a similar way.
6. Oxidative Stress and Amyloid-Beta Aggregation: Some bacterial infections have been associated with increased oxidative stress, which can exacerbate the aggregation of Aβ and tau, key pathological features in AD [16]. By increasing oxidative stress, a K. pneumoniae infection could indirectly encourage the accumulation of these proteins in the brain, contributing to neurodegeneration.

In sum, while direct evidence linking K. pneumoniae to Alzheimer’s disease is still under investigation, the pathways of inflammation, immune activation, BBB disruption, and potential neuroinvasion provide a plausible basis for how recurrent or chronic K. pneumoniae infections could increase Alzheimer’s risk, especially in older adults or individuals with compromised immune systems.

## Data Availability

All data produced are available online at UK Biobank https://www.ukbiobank.ac.uk/

## Acknowledgment

Graphical abstract created in https://BioRender.com

